# Nanopore target sequencing for accurate and comprehensive detection of SARS-CoV-2 and other respiratory viruses

**DOI:** 10.1101/2020.03.04.20029538

**Authors:** Ming Wang, Aisi Fu, Ben Hu, Yongqing Tong, Ran Liu, Jiashuang Gu, Jianghao Liu, Wen Jiang, Gaigai Shen, Wanxu Zhao, Dong Men, Zixin Deng, Lilei Yu, Yan Li, Tiangang Liu

**Affiliations:** Department of Clinical Laboratory, Renmin Hospital of Wuhan University, Wuhan, 430060, China; Key Laboratory of Combinatorial Biosynthesis and Drug Discovery, Ministry of Education and Wuhan University School of Pharmaceutical Sciences, Wuhan, 430071, China; Wuhan Dgensee Clinical Laboratory Co., Ltd., Wuhan, 430075, China; Wuhan Institute of Virology, Chinese Academy of Sciences, Wuhan, Hubei Province, P.R. China; State Key Laboratory of Microbial Metabolism, Joint International Research Laboratory of Metabolic & Developmental Sciences, and School of Life Sciences and Biotechnology, Shanghai Jiao Tong University, Shanghai, 200030, China; Hubei Engineering Laboratory for Synthetic Microbiology, Wuhan Institute of Biotechnology, Wuhan, 430075, China; Department of Internal Medicine, Renmin Hospital of Wuhan University. Wuhan, 430060, China

## Abstract

The ongoing novel coronavirus pneumonia COVID-19 outbreak in Wuhan, China, has engendered numerous cases of infection and death. COVID-19 diagnosis relies upon nucleic acid detection; however, current recommended methods exhibit high false-negative rates, low sensitivity, and cannot identify other respiratory virus infections, thereby resulting patient misdiagnosis and impeding epidemic containment. Combining the advantages of target amplification and long-read, real-time nanopore sequencing, we developed nanopore target sequencing (NTS) to detect SARS- CoV-2 and other respiratory viruses simultaneously within 6–10 h. Parallel testing with approved qPCR kits of SARS-CoV-2 and NTS using 61 nucleic acid samples from suspected COVID-19 cases confirmed that NTS identified more infected patients as positive, and could also monitor for mutated nucleic acid sequence or other respiratory virus infection in the test sample. NTS is thus suitable for contemporary COVID-19 diagnosis; moreover, this platform can be further extended for diagnosing other viruses or pathogens.

## Introduction

An ongoing novel coronavirus pneumonia (COVID-19) outbreak originating in Wuhan, China in December 2019 has subsequently spread across China and worldwide, resulting in numerous cases of infection and death^1^. Usually, COVID-19 has an incubation period of 2–7 days^2^ with no obvious symptoms, during which time the virus can spread from infected to uninfected individuals.Therefore, early accurate diagnosis and isolation of patients is key to controlling the epidemic.

Nucleic acid detection is the golden standard for COVID-19 diagnosis. Real-time reverse transcription-polymerase chain reaction (qPCR) is the most recommend testing method for detecting the causative virus, SARS-CoV-2^3^. qPCR is specific, rapid, and economic, but cannot precisely analyze amplified gene fragment nucleic acid sequences; thus, positive infection is confirmed by monitoring one or two sites (depending on manufacturer guidelines). However, qPCR exhibits high false-negative rates and low sensitivity in clinical application^4^, with only 30–50% positive detection ratio. False-negatives facilitate epidemic spread through delayed patient isolation and treatment, and patients mistakenly considered uninfected or cured following misdiagnosed treatment results. Another recommend detection method, sequencing, is widely applied for pathogen identification and monitoring virus evolution^5, 6^ including SARS-CoV-2^7^, but requires expensive equipment, operator expertise, and > 24 h turnaround time, rendering it unsuitable for the current crisis.

Several intelligent methods for RNA virus detection have developed including combining toehold switch sensors^8^, which can bind to and sense virtually any RNA sequence, with paper-based cell-free protein synthesis for Ebola and Zika virus detection^9, 10^, and the SHERLOCK method based on CRISPR/Cas13a for Zika or Dengue virus detection^11^. A rapid SHERLOCK method with visual results can detect SARS-CoV-2^12^ and toehold switch biosensors could theoretically be developed for rapid and high-throughput SARS-CoV-2 detection. However, the requirement of specific RNA regions as targets may negatively affect detection rates because target region mutation may limit target availability.

Moreover, pneumonia and fever may be caused by other respiratory viruses^13^. Cross-infection during the diagnosis process both spreads SARS-CoV-2 and subjects COVID-19 patients to other respiratory viruses. In severe cases, comprehensive analysis of infecting viruses is necessary.Therefore, a rapid, accurate, and comprehensive detection method is needed to inform clinical treatment and control cross-infection to reduce mortality.

Currently, COVID-19 infection and death rates in Hubei province are the highest in China. Being located at the center of this epidemic, we developed a nanopore target sequencing (NTS) method combining the advantages of target amplification and long-read, real-time nanopore sequencing for detecting SARS-CoV-2 with higher sensitivity than standard qPCR, simultaneously with other common respiratory viruses and mutated nucleic acid sequence within 6–10 h.

## Results

### NTS design for SARS-CoV-2 detection

NTS is based upon amplification of 11 virulence-related and specific gene fragments (*orf1ab*) of SARS-CoV-2 using a primer panel developed in-house, followed by sequencing the amplified fragments on a nanopore platform. To enhance sensitivity, we focused on virulence-related genes as targets without limitation to the sites currently recommended by Chinese or American Centers for Disease Control (CDC) in qPCR methods (Fig. 1). Because this method can precisely determine nucleic acid sequences, positive infection can be confirmed by analyzing output sequence identity, coverage, and read number.

**Fig. 1.**
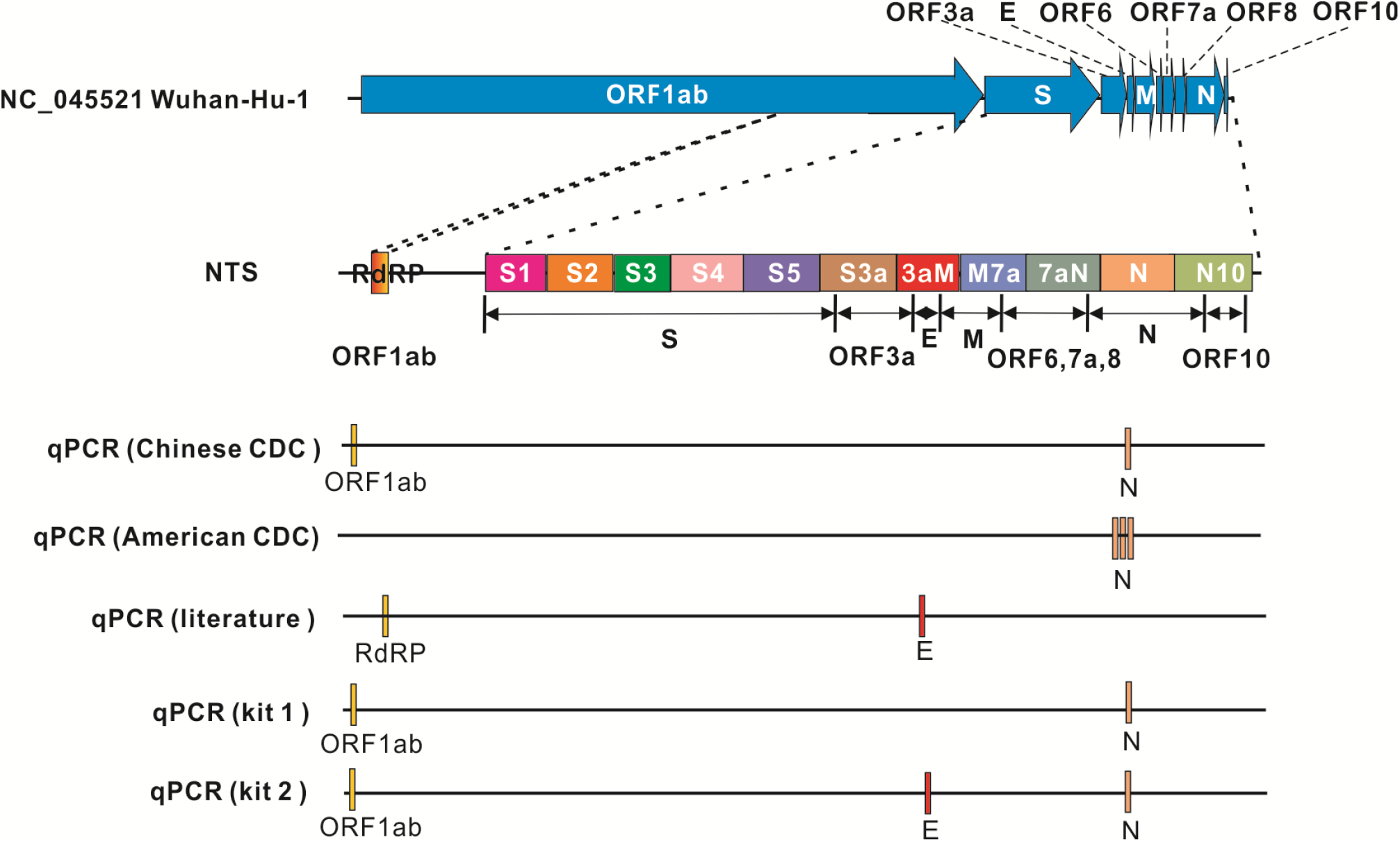
Amplification targets of the NTS and qPCR method. NTS detected 12 fragments including ORF1ab and virulence factor-encoding regions. For qPCR, the Chinese CDC recommends *Orf1ab* and *N* sites as targets,^36^ the United States CDC recommends three target sites in the *N* gene,^37^ and Corman *et al* (2020) recommend RNA-dependent RNA polymerase (RdRP) in *orf1ab* and *E* sites as the targets. Kit 1 is a cFDA-approved kit with two target sites used in this study; kit 2 is a cFDA-approved kit with three target sites used in this study.

To realize detection of pivotal SARS-CoV-2 virulence genes, we focused on the virulence region (genome bp 21,563–29,674; NC_045512.2), encoding S (1273 amino acids; AA), ORF3a (275 AA), E (75 AA), M (222 AA), ORF6 (61 AA), ORF7a (121 AA), ORF8 (121 AA), N (419 AA), and ORF10 (38 AA) proteins. We also considered the RNA-dependent RNA polymerase (RdRP) region in *orf1ab* (Fig. 1). For the virulence regions, 11 fragments of 600–950 bp were designed as targets, fully covering the 9,115 bp region (Fig.1), amplified by 22 specific primers designed considering primer-primer interaction and annealing temperature, and potential non-specific binding to human and common bacterium and fungi genomes. To improve the sensitivity *orf1ab* region amplification, we designed two pairs of nested primers to amplify 300–500 bp regions to avoid amplification failures owing to site mutation. Finally, the 26 primers were combined to develop the SARS-CoV-2 primer panel (Supplementary Table 1).

For sequencing, we chose a nanopore platform that could sequence long nucleic acid fragments and simultaneously analyze the data-output in real-time. This allowed confirmation of SARS-CoV-2 infection within a few minutes after sequencing by mapping the sequence reads to the SARS-CoV-2 genome and analysis of output sequence identity, coverage, and read number. Moreover, the accurate nucleic acid sequence generated using our pipeline could indicate whether the virulence-related genes were mutated during virus spreading, thereby rapidly providing information for subsequent epidemiological analysis. Additionally, as the MinION nanopore sequencer is portable, NTS is also suitable for front-line clinics.

### NTS results interpretation and limit of detection (LoD)

To test the SARS-CoV-2 detection efficiency by NTS, we used standard plasmids harboring COVID-19 virus *S* and *N* genes to simulate SARS-CoV-2. Standard plasmids were individually spiked into background cDNA samples (cDNA reverse-transcribed from an uninfected respiratory flora throat swab) at 10, 100, 500, 1000, and 3000 copies/mL. NTS for all test samples was performed on one MinION sequencer chip. Sequence data were evaluated at regular intervals using our in-house bioinformatics pipeline. By mapping output reads on the SARS-CoV-2 genome, all reads with high identity were calculated for each plasmid concentration. For 10 min and 1 h sequencing data, reads mapped to SARS-CoV-2 significantly differed from those of negative controls in all replicates at concentrations ranging from 3000 to 500 (Fig. 2a), and 3000 and 10 (Fig. 2b) copies/mL, respectively. This result confirmed that high-copy samples could rapidly yield sufficient valid sequencing data for diagnosis, and by extending the sequencing time, valid sequencing data could also be obtained from low-copy samples. Notably, as more sequencing data could be achieved with additional sequencing time (Supplementary Fig. 1) and clinical samples may exhibit higher complexity, thus, 10 min (for quick detection) and 4 h (for final evaluation) sequencing times were used in subsequent evaluation of NTS in clinical samples.

**Fig. 2.**
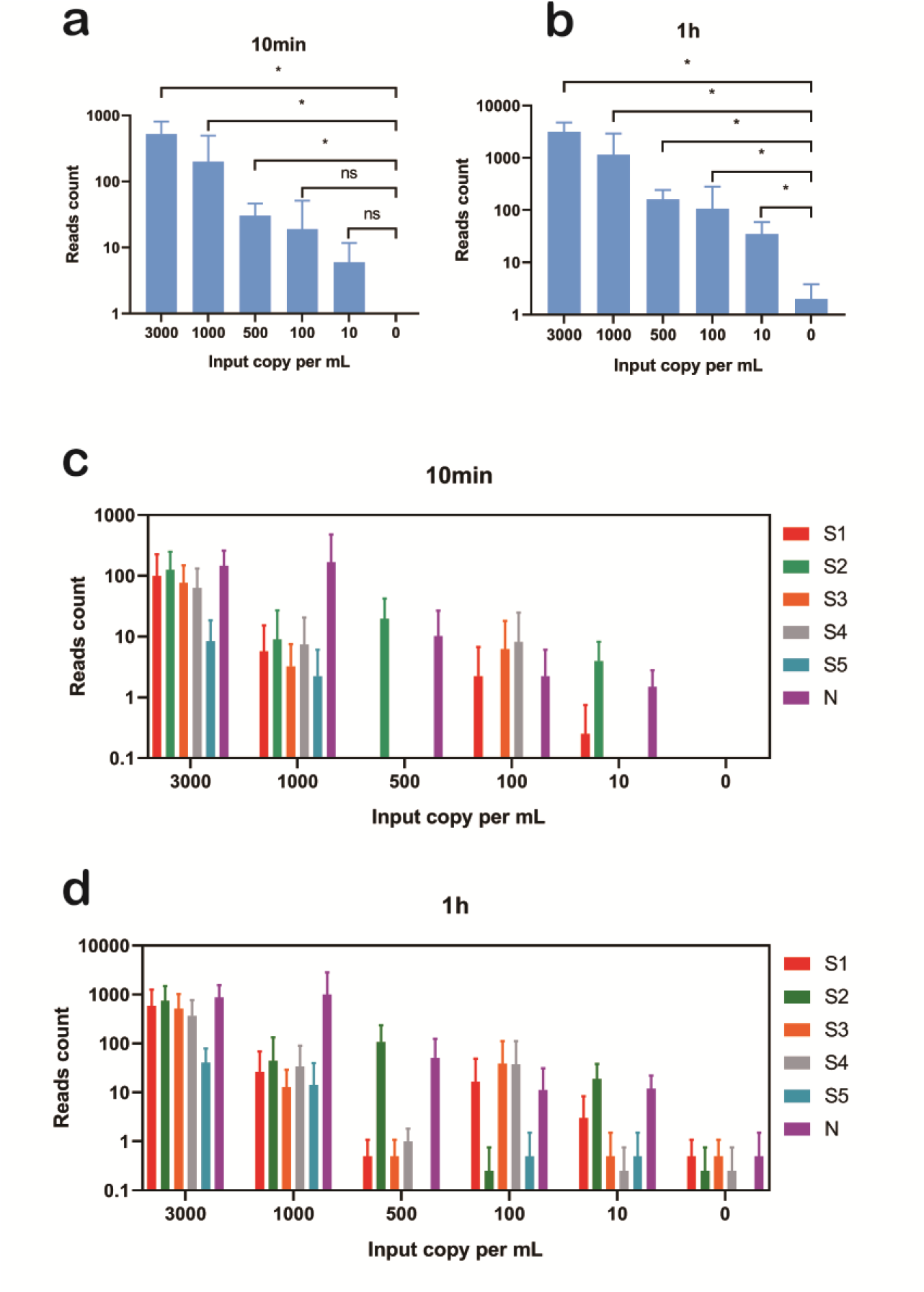
Performance verification test of NTS for detecting SARS-CoV-2 using standard synthetic *S* and *N* genes. Comparison of all SARS-CoV-2 reads detected by NTS in replicates with different concentrations and negative controls using 10 min (**a**) or 1 h (**b**) sequencing data. Read counts mapped to each target region of the SARS-CoV-2 genome in replicates with different concentrations with 10 min (**c**) to 1 h (**d**) sequencing data. Two-tailed Student t-test (for normal distribution samples) or Mann–Whitney U-test (for non-normal distribution samples): ns, not significant, \**P* < 0.05; bars represent the means ± SD.

Evaluation of the target distribution of these valid data revealed that in higher copies samples (1000 and 3000 copies/mL), all targeted regions could be detected (Fig. 2c, d). However, in lower viral concentration samples (from 10 to 500 copies/mL), some targeted regions were lost (i.e., no reads mapped; Fig. 2c, d), indicating that for low-quality or low-abundance samples, comprehensive fragment amplification is difficult. Therefore, for accurate results, NTS cannot label a sample as positive for infection by monitoring only one or two sites, as is customary for qPCR; rather, the results from all target regions should be considered.

Accordingly, we determined a scoring rule by referring to previous judgment rules^14-16^. Firstly, we counted the number of output reads with high identity to the SARS-CoV-2 genome, indicative of high credibility for identification as SARS-CoV-2. By calculating the ratio of the counted valid read numbers of the test sample to those of the negative control (with “0” in the negative control calculated as “1”), we defined that a ratio of ≥10 indicates a positive result for that fragment, scoring 1; ≥3 to 10 fold is inconclusive, scoring 0.4; and <3 is negative, scoring 0. Scores were summed to obtain the NTS score. We considered that a sample in which at least 50% fragments (6 fragments) are inconclusive or 2 fragments are positive (comparable to qPCR results) could be defined as a positive infected sample (e.g., NTS score >2.4); 3–6 inconclusive fragments or 1 positive fragment indicated a highly suspect (inconclusive) sample (e.g., NTS score of 1.2–2.4); and < 3 inconclusive or no positive fragments could be defined as negative sample (NTS score <1.2).

To determine the NTS LoD, we used the defined rules to evaluate each replicate in the simulated test. As the standard plasmids contain only 6 designed fragments (half of 12 designed fragments for SARS-CoV-2), we defined the scoring as NTS score >1.2 indicates positive detection, 0.6–1.2 is inconclusive, and < 0.6 reflects negative detection. We calculated the score of the lowest concentration (10 copies) at different times according to this scoring method and judged the positive detection rate. The results (Supplementary table 2) showed that 3/4 of the 10 copies of the standard plasmids can be judged positive from 1 h. This result is consistent with the significant comparation (Fig. 2b), that the data for 10 copies standard plasmids is also significantly different from the negative control from 1h. This result shows that our scoring system is reliable for evaluating NTS test results, and the LoD (3/4 replicates positive) was determined as 10 copies/mL with 1h sequencing data (1,372 to 43,967 reads per sample in a run with 24 samples).

### SARS-CoV-2 detection using qPCR vs NTS

We performed clinical sample testing at the first-line hospital in Wuhan as soon as NTS method was established (Fig. 3). To verify NTS sensitivity, we evaluated 45 nasopharyngeal swab samples from outpatients with suspected COVID-19 early in the epidemic. On February 6 and 7, 2020, we parallel tested these 45 samples in two batches using NTS (two chips) and qPCR (kit 2; Fig. 1). The 4 h sequencing output data (Fig. 4a) revealed that all 19 samples defined as positive by qPCR were recognized SARS-CoV-2-infected by NTS, indicating good inter-test concordance. Among 15 qPCR-inconclusive samples, 11 were recognized as SARS-CoV-2-infected, 3 as negative, and 1 inconclusive by NTS. Among 11 qPCR-negative samples, 4 were recognized SARS-CoV-2-infected, 4 as inconclusive, and 3 as negative by NTS. Overall, NTS identified a total of 34 positive samples in 45 suspected samples, 15 more than qPCR. Evaluation of output data after 10 min sequencing (Supplementary Fig. 2) revealed that 21 of 45 suspected samples were recognized as SARS-CoV-2-infected by NTS. For these samples, the 10 min and 4 h results were comparable, indicating that NTS could rapidly detect many positive samples.

**Fig. 3.**
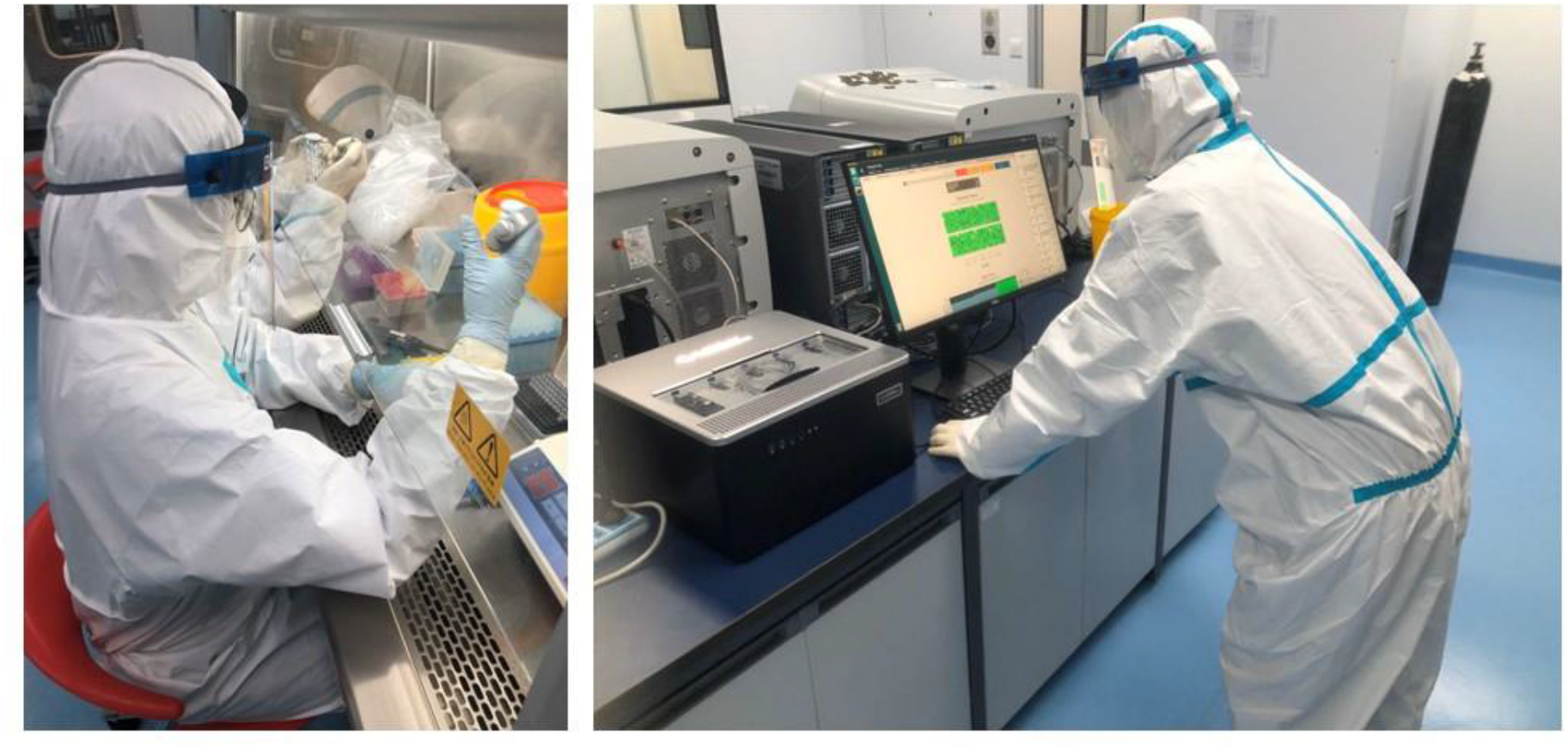
NTS testing in a front-line hospital in Wuhan.

**Fig. 4.**
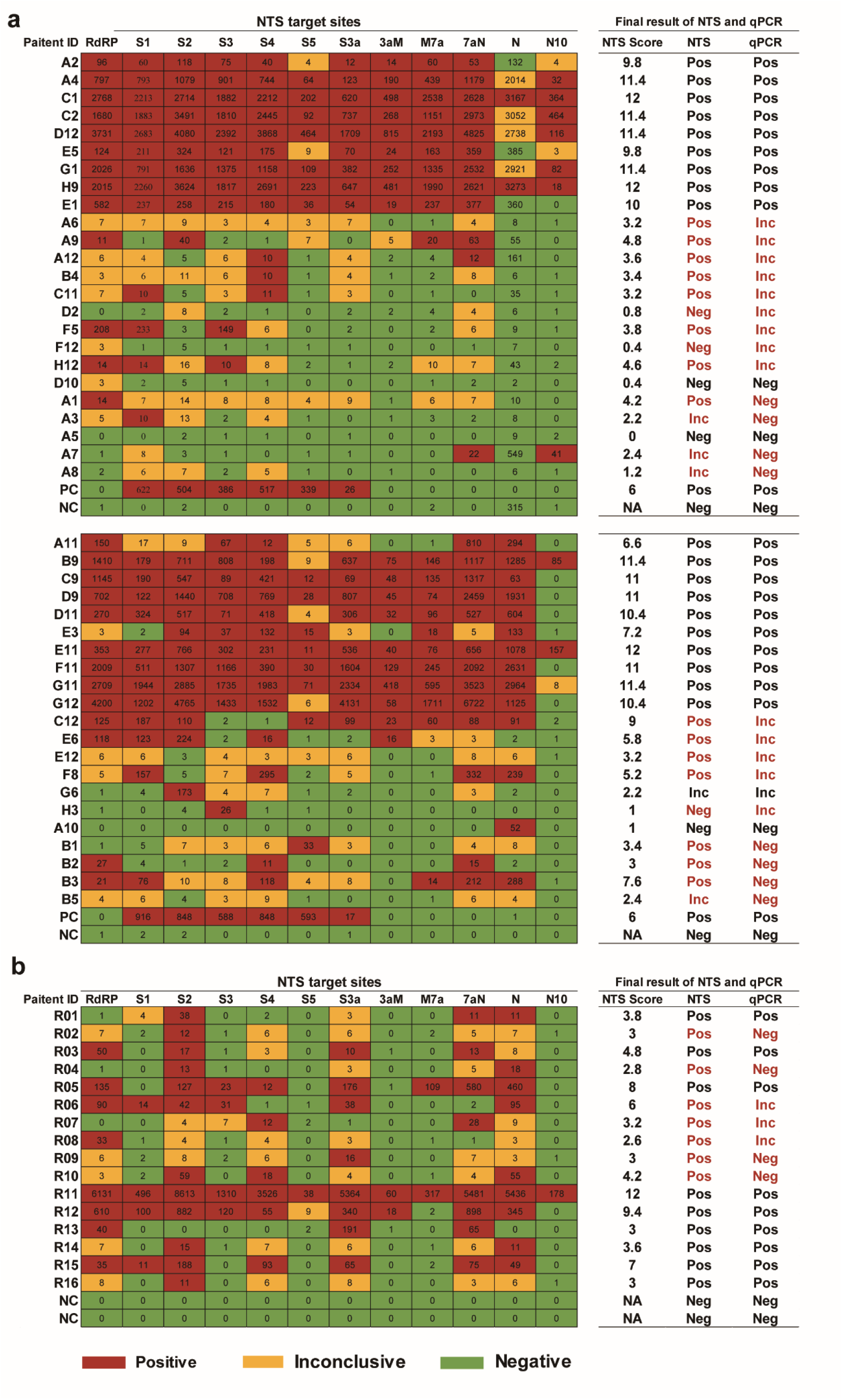
Comparison of 61 nucleic acid sequences from clinical samples obtained using NTS (4 h) and qPCR. **a**, Comparison of 45 nucleic acid sequences from samples of patients with suspected COVID-19 obtained using NTS and qPCR (kit 2). **b**, Comparison of 16 nucleic acid sequences from patients with confirmed disease obtained using NTS and qPCR (kit1). The numbers in the table on the left represent the number of mapped reads according to our rules. PC: positive control. The plasmid harboring an *S* gene was used as a positive control in NTS testing; a positive sample in the kit served as a positive control in qPCR testing. NC: negative control. TE buffer was used as a negative control in NTS testing; H2O in the kit served as a positive control in qPCR testing. Pos: positive. Inc: inconclusive. Neg: Negative.

However, as the tested 45 samples were from early outpatients without detailed records, suitable clinical data, such as chest computed tomographic scans, were not available to support the results. Therefore, we next evaluated samples retained from hospitalized patients with confirmed COVID-19 subjected to qPCR testing (kit 1, Fig. 1) on February 11 and 12, 2020. We randomly selected 16 samples for NTS testing on February 20, 2020. 4 h sequencing (Fig. 4b) identified all 16 positive samples, whereas only 9 samples were positive by qPCR. At the time of this writing, among these 7 samples that qPCR negative or inconclusive, electronic records indicated that subsequent qPCR testing of 4 of these 7 patients revealed two (R04 and R09) as positive whereas two (R06 and R07) remained inconclusive. This result confirmed that NTS could identify more positive cases than qPCR. Three positive samples were identified by 10 min sequencing (Supplementary Fig. 2), indicating that NTS could rapidly detect positive samples with high concentration of virus.

Evaluation of the positive target distribution for each sample (Fig. 4) showed that samples positive by both NTS and qPCR appeared to have higher nucleic acid quality or abundance, because NTS yielded more positive fragments. For qPCR-inconclusive samples, NTS yielded few, scattered positive target fragments, suggesting that low sample nucleic acid quality or abundance rendered it difficult to draw clear conclusions by qPCR based on evaluation of only two sites. Moreover, contamination of individual viral fragments did not affect NTS results. For example, although the negative control of the first chip in Fig. 4 appears to have been contaminated with a fragment containing the *N* gene, we could exclude the contamination using a high threshold, and/or base the final conclusions on the 11 remaining sites. However, negative control contamination in qPCR would invalidate the results of the whole experimental batch.

### SARS-CoV-2 mutation analysis

Mutation screening of 19 samples from outpatients indicated as infection-positive by both NTS and qPCR identified single nucleotide mutations at seven sites in four samples (Table 1), three of which (S_519 of C1, N_822 of C2, and S_2472 of E3) harbored silent mutations. One variant (ORF8_251: T→C), encoding an AA change from Leu to Ser, was identified in the three samples. The ORF8_184 mutation in sample E3 also reflected a Val to Leu missense mutation. Comparison with the 67 complete SARS-CoV-2 genomes reported in the GISAID database prior to February 8, 2020 revealed that ORF8_251 contained C in 20, T in 48, and Y in 1 genome, indicating its diversity in different strains. Additionally, single genomes contained C or S at ORF8_184 whereas the remaining 67 had G, indicating that despite some inter-strain diversity, G→C transversion was rare. The remaining silent variants were not identified in the GISAID database, suggesting that the virus may harbor mutations as yet uncharacterized by existing genome-wide sequencing methods.

**Table 1.** Variations of SARS-CoV-2 detected by NTS.

| Sample | Site<br>(Gene_position) | Base change | Base change present<br>in the genome<sup>a</sup> | Amino acid change |
| --- | --- | --- | --- | --- |
| C1 | S_519 | G→A | 0 | NC |
| C2 | ORF8_251 | T→C | 20 | Leu→Ser |
| C2 | N_822 | C→T | 0 | NC |
| E3 | S_2472 | C→T | 0 | NC |
| E3 | ORF8_251 | T→C | 20 | Leu→Ser |
| E3 | ORF8_184 | G→C | 1 | Val→Leu |
| E5 | ORF8_251 | T→C | 20 | Leu→Ser |

<sup>a</sup> The number indicates the count of genomes in which the base change appeared as reported in the GISAID database prior to February 8, 2020. NC: no change.

### NTS panel for respiratory virus identification

The inability of current clinically utilized SARS-CoV-2 qPCR kits to identify the species of co-infecting viruses combined with the high false-negative rate of qPCR compromises early patient triage, resulting in wasted urgent medical resources and enhancing potential cross-contamination during the diagnosis process. Distinguishing different types of respiratory viral infections has attracted worldwide attention.

To extend the scope of NTS-based virus detection, we designed a respiratory virus primer panel for amplification of 10 respiratory viruses including bocavirus, rhinovirus, human metapneumovirus, respiratory syncytial virus, coronavirus, adenovirus, parainfluenza virus, influenza A virus, influenza B virus, and influenza C virus. We collected target gene candidates utilized for virus identification in the literature, then collected all complete and partial target gene sequences for these viruses available at GenBank (through November 1, 2019). Though multiple nucleic acid sequence alignment of each gene, the conserved regions were chosen as candidate regions for amplification. Using similar constraints as for SARS-CoV-2 target region selection, we chose 20 target amplification regions (300–800 bp) for these 10 respiratory viruses (Supplementary Table 3) capable of accurately distinguishing virus in addition to identifying virus species. We designed 59 primers including some nested primers for amplification of these regions, comprising the respiratory virus primer panel (Supplementary Table 4).

To verify the performance of this panel in NTS, we selected five virus-positive samples (influenza A virus, influenza B virus, parainfluenza, respiratory syncytial virus, and rhinovirus), the viruses in the clinical throat swabs were previously confirmed using a China Food and Drug Administration (cFDA) approved kit (Health Gene Technologies, China) based on multiplex PCR and capillary electrophoresis analysis. The five samples were mixed to create a mock virus community and used to test the NTS virus detection capacity. NTS 10 min sequencing data (Supplementary Table 5) successfully detected four of five viruses (influenza A virus, influenza B virus, respiratory syncytial virus, and rhinovirus); the remaining one virus with lower load could be detected with 2 h sequencing data, confirming the suitability of NTS with the respiratory virus primer panel for identification of other respiratory viruses.

To verify the ability of NTS to detect SARS-CoV-2 and 10 kinds of respiratory viruses in a single assay, 13 of the 45 suspected COVID-19 outpatient samples were subjected to simultaneous detection analysis. Five replications of the plasmid containing the SARS-CoV-2 *S* and *N* genes served as the positive control and Tris-EDTA (TE) buffer was used as the negative control (in duplicate). For each sample, cDNA samples were separately amplified using the respiratory virus and the SARS-CoV-2 primer panels, then all amplified fragments were pooled. After the addition of barcodes, amplified fragments from all 20 samples (13 cases, 7 controls) were subjected nanopore sequencing on one chip. Analysis of the results (Table 2) revealed that E11 was co-infected by influenza A virus H3N2 and SARS-CoV-2.

**Table 2.** Results of NTS for detecting SARS-CoV-2 and common respiratory viruses.

| Patients ID | NTS result<br>(SARS-CoV-2) | NTS result<br>(common respiratory viruses) |
| --- | --- | --- |
| F11 | Positive | ND |
| E11 | Positive | human influenza A virus H3N2 |
| A11 | Positive | ND |
| B9 | Positive | ND |
| C9 | Positive | ND |
| D9 | Positive | ND |
| D11 | Positive | ND |
| C12 | Positive | ND |
| E6 | Positive | ND |
| B3 | Positive | ND |
| E12 | Negative | ND |
| G6 | Inconclusive | ND |
| B1 | Inconclusive | ND |
| Positive control | Positive | ND |
| Positive control | Positive | ND |
| Positive control | Positive | ND |
| Positive control | Positive | ND |
| Positive control | Positive | ND |
| Negative control | ND | ND |
| Negative control | ND | ND |

ND: not detected

## Discussion

Herein, we developed an NTS method able to simultaneously detect SARS-CoV-2 and 10 additional types of respiratory viruses within 6–10 h, at LoD of 10 copies/mL with at least 1 h sequencing data. The detection region of SARS-CoV-2 was composed of 12 fragments covering nearly 10 kb of the genome, resulting in markedly higher sensitivity and accuracy than those of qPCR kits currently in clinical use. Notably, 22 of 61 suspected COVID-19 samples that were negative or inconclusive by qPCR testing were identified as positive by NTS. Moreover, NTS enabled the detection of virus mutations; in particular, we detected a nucleotide mutation in SARS-CoV-2 that was undetected in the genomic data in the current GISAID database. Although this was a silent mutation, its presence suggests that the virus may have undergone mutation during the spreading process. Additionally, NTS was verified as capable of detecting all five pre-added respiratory viruses in a single detection. This method also detected a co-infected case (SARS-CoV-2 and human influenza A virus H3N2) using a clinical specimen, illustrating the ability of NTS to detect and distinguish respiratory viruses. Together, our findings indicate that NTS is highly suitable for the detection and variation monitoring of current COVID-19 epidemics, directly from clinical samples with same-day turnaround of results.

At the time of this writing, the COVID-19 epidemic remains very severe. Accurate, rapid, and comprehensive nucleic acid detection methods are needed to allow patients with suspected infection to be isolated and treated as soon as possible, and to accurately confirm whether the patient is cured, to prevent continued epidemic spread caused by misdiagnosis. The LoD of NTS was shown to be as low as 10 copies/mL, rendering it 100-fold more sensitive than qPCR (e.g., some kits describe LoDs of 1000 copies/mL) and thus likely to decrease the high false-negative rate plaguing current detection methods. In addition, the detection of co-infection may allow the prevention of disease progression from mild to severe or might be useful to inform clinical treatment. Overall, NTS combines sensitivity, broad detection range, same-day rapid turnaround time, variation monitoring, and low cost (compared with whole-genome sequencing), making it the most suitable method for the detection of suspected viral infections that cannot be effectively diagnosed by other methods. Moreover, the MinION, the smallest Oxford Nanopore sequencer, is smaller than a cellphone; when equipped with a laptop computer for data processing, it thus allows rapid performance of NTS in various environments with low equipment cost. For data analysis, cloud analysis may also be introduced for high-throughput detection^17, 18^.

Several limitations of the current NTS method should be noted. Because the designed amplified fragments are 300–950 bp in length, which constitute suitable lengths for detection by a nanopore platform as nucleic acid fragments < 200 bp cannot be readily detected^19, 20^, thereby, the sensitivity of NTS for detecting target COVID-19 fragments in highly degraded nucleic acids may be hampered. Additionally, although the turnaround time of NTS is longer than that of qPCR or other possible nucleic acid detection methods (e.g., SHERLOCK^12^), 6–10 h is considered acceptable for clinical use; moreover, NTS is already the fastest strategy based on sequencing methods to date and can detect variations directly from clinical samples. Whereas the detection throughput of NTS is not high at present, the NTS method can be integrated into widely used automated or semi-automated platforms to improve the detection throughput in the future^21-23^. In addition, because PCR is included in NTS, processes involving opening the lid of the PCR tubes may cause mutual contamination between samples^24, 25^. However, this situation also is inevitable in current nucleic acid detection methods (e.g., qPCR) or other nucleic acid detection schemes (e.g., SHERLOCK^11, 12^ or toehold switch biosensor^9, 10^) that also involve PCR. The introduction of integration systems or sealed devices such as microfluidics may avoid this situation^26, 27^. At present, our processes of sequencing data analysis and interpretation of results are not mature; nevertheless, as the number of test samples increases, additional test results will be collected and the process continuously optimized to obtain more accurate results.

Notably, the comparison of NTS and qPCR results indicated a high false-negative rate in the latter. This result highlights the need for extreme vigilance, as patient misdiagnosis (including patients admitted and discharged) will lead to spread of the epidemic and greater public health threat. Suspected or negative results reported by the current qPCR methods should be subjected to a more accurate method for secondary confirmation; for this, we consider NTS as the most recommended solution currently available. The situation of co-infection, which has been reported in our previous study^13^, also warrants continued attention. Based on the current centralized treatment strategy, the lack of screening for multiple viruses may lead to large-scale cross-contamination and confound clinical diagnosis and treatment. Alternatively, NTS represents and effective strategy that can rapid and accurate distinguish SARS-CoV-2 and multiple respiratory viruses at both the species and subtype level, and could be applied as a spot check in centralized clinics. Finally, the NTS method for respiratory virus detection might be extended to detect more viruses and other pathogens through the design of additional primer panels.

## Methods

### Primer panel design for SARS-CoV-2

The SARS-CoV-2 primer panel was designed to simultaneously detect virus virulence- and infection-related genes and variants thereof. The 21,563– 29,674 bp genome region, containing the genes encoding S, ORF3a, E, M, ORF6, ORF7a, ORF8, N, and ORF10, was selected as a template to design a series of end-to-end primers. The region encoding ORF1ab was selected as a template to design a nested primer for higher sensitivity detection of SARS-CoV-2. All primers were designed using online primer-blast (https://www.ncbi.nlm.nih.c/tools/primer-blast/) and the specificity of all primers was verified against *Homo sapiens*, fungi, and bacteria. Finally, we downloaded and selected *N, S, rdrp*, and *E* gene sequences of SARS-related viruses available at GenBank through January 1st, 2020 (accession NC_045512). Multiple sequence alignment of SARS-CoV-2 against SARS-related viruses was performed using Clustal W (version 1.83) for each gene individually and the alignment was used for in-silico evaluation of specificity of the designed primers to SARS-CoV-2. All the specific primers were collected to form the SARS-CoV-2 primer panel.

### Primers panel design for 10 kinds of respiratory virus detection

The target genes for each virus were selected based on previous literature retrieval and all complete and partial gene sequences available in GenBank through November 1, 2019 were downloaded. The list for each target gene was manually checked and artificial sequences (e.g., lab-derived, synthetic) along with sequence duplicates was removed, resulting in a final list. Multiple sequence alignment was performed using Clustal W (version 1.83) for each gene individually and the variation rate of each base was calculated using an in-house pipeline. The final primers for each virus were manually selected following the previous metrics^28^ for multiplex PCR design with an expected amplicon length range from 300 to 800 bp.

### LoD of the NTS test

Individual NTS libraries were prepared from a virus-negative nasopharyngeal swab spiked with plasmids containing synthetic *S* and *N* genes of COVID-19 at concentrations of 0, 10, 100, 500, 1,000, and 3,000 copies/mL, with four replicates at each concentration. The NTS libraries were prepared as described above and sequenced using MinION for 10 min, 30 min, 1 h, 2 h, and 4 h. The sequencing data were processed as described for virus identification. The LoD was determined when the concentration of reads mapped to COVID-19 was significantly higher than that for the negative control in 3/4 replicates.

### NTS detection method

The targeted genes were amplified using the SARS-CoV-2 or 10 respiratory virus primer panel in a 20 μL reaction system with 5 μL total nucleic acid, 5 μL primer (10 μM), and 10 μL 2× Phusion U Multiplex PCR Master Mix (Thermo Fisher, USA)^29, 30^. Amplification was performed in a C1000 Thermocycler (Bio-Rad, USA) using the following procedure: 1 cycle at 94 °C for 3 min and 30 cycles at 95 °C for 10 s, 55 °C for 50 s, and 68 °C for 5s, followed by a final elongation step at 68 °C for 5 min. The product of the first-step was purified with 0.8× AMpure beads (Beckman Coulter, USA) and eluted in 10 μL Tris-EDTA (TE) buffer. Then, 5 μL eluate was used for second-step PCR with 5 μL barcoded primer (10 μM) and 10 μL 2× Phusion U Multiplex PCR Master Mix. The barcode sequence was from the Nanopore PCR barcode kit (EXP-PBC096; UK) and all primer oligos and full-length *S* and *N* gene fragments were synthesized by Genscript (China). The products of second-step PCR from the different samples were pooled with equal masses. TE buffer was assayed in each batch as a negative control. Sequencing libraries were constructed using the 1D Ligation Kit (SQK-LSK109; Oxford Nanopore, UK) and sequenced using Oxford Nanopore MinION or GridION.

### Nanopore sequencing data processing

Basecalling and quality assessment for MinION sequencing data were performed using high accuracy mode in Guppy (v. 3.1.5) software; for GridION, the process was conducted using MinKNOW (v. 3.6.5) integrated in the instrument. Sequencing reads with low quality and undesired length were discarded. Then, Porechop^31^ (v. 0.2.4) was used for adaptor trimming and barcode demultiplexing for retained reads.

### Mapping tool and mapping database

BLASTn^32^ (v. 2.9.0+) was used to map the reads of each sample against the virus genome reference database. All virus genomic sequences were downloaded from NCBI Refseq FTP and the SARS-CoV-2 genome sequence was added to the BLAST database subsequently. The taxonomy of each read was assigned according to the taxonomic information of the mapped subject sequence.

### Sequence correction and candidate mutation calling

Sequence correction was performed using medaka^33^ (v. 0.10.5), which is a tool to create a consensus sequence of nanopore sequencing data. For each target sequencing region, 30 consensus sequences were generated using medaka’s default settings through the medaka_consensus program. Subsequently, the consensus sequences were aligned to the reference sequence of target sequencing regions using the multiple sequence alignment tool ClustalW^34^ (version 1.83). The variants within certainty regions (except sequence homopolymeric regions and primer binding sites)^35^ and with appropriate coverage (covered by at least 90% consensus sequences and at least 50% uncorrected reads) were accepted as candidate nucleotide mutations.

### Interpretation of NTS results

The sequenced data were obtained at regular intervals after sequencing, then filtered to obtain valid reads. For determining whether the target was positive, interpretation was performed using the previous rule with modification^14-16^. In brief, if the read matches the design fragment, the read will be counted. The mapping score was determined as 1, 0.4, or 0 when the ratio of count number in the sample to the negative control of each target was > 10, between 3–10, or < 3. The total mapping score of each target was summed and samples with > 2.4 total mapping score were defined as positive for SARS-CoV-2 infection; 1.2 to 2.4 total mapping score indicated an inconclusive result, and < 1.2 total mapping score was considered to indicate negative for infection. For determination of the other 10 kinds of common respiratory virus, a sample was considered positive for the virus if positive for at least one designed site, otherwise it was negative.

### Total nucleic acid extraction from clinical specimens

Clinical throat swab specimens were collected in 10 mL of Viral Transport Medium (Becton Dickinson, USA) from 45 suspected COVID-19 outpatients, and 16 hospitalized patients with COVID-19 at Renming Hospital of Wuhan University in Wuhan during February 2020. All throat swabs were sent to a clinical laboratory and processed immediately. Swabs were vortexed in 1 mL of TE buffer and centrifuged at 20,000 × *g* for 10 min. The supernatant was removed and 200 μL of the specimen was retained for total nucleic acid extraction. Total nucleic acid was extracted from 200 μL of pre-treated samples using the Sansure SUPRall DNA Extraction Kit (Changsha, China) following the manufacturer’s instructions. Extracted total nucleic acid was stored at 70 °C until qPCR or NTS testing.

### qPCR for confirmation of SARS-Cov-2 infection

The total isolated nucleic acid was used for qPCR assay following the manufacturer’s instructions. Briefly, qPCR was carried out in a 25 μL reaction system using a novel coronavirus qPCR kit (kit 1, Huirui, China) with 5 μL total nucleic acid, or 20 μL reaction system using the 2019-nCoV qPCR kit (kit 2, BioGerm, China) with 5 μL total nucleic acid. For kit 1, amplification was performed using a Quantstudio Dx Real-time PCR system (Thermo Fisher, USA) with the following procedure: 1 cycle at 50 °C for 15 min and 95 °C for 5 min, and 35 cycles at 95 °C for 10 s, 55 °C for 40 s. The FAM and ROX fluorescence channels were used to detect *Orf1ab* and *N* gene, respectively. Successful amplification of both genes and Ct value ≤35 was recognized as positive for SARS-CoV-2 infection; Ct value between 35.2 to 39.2 was recognized as inconclusive, and one of the genes being undetected or Ct ≥ 39.2 was recognized as negative. For kit 2, amplification was performed in a CFX96 Thermocycler (Bio-Rad) using the following procedure: 1 cycle at 50 °C for 10 min and 95 °C for 5 min, and 35 cycles at 95 °C for 10 s, 55 °C for 40 s. The FAM, HEX, and CY5 fluorescence channels were used to detect *Orf1ab, E*, and *N* genes, respectively. This kit only utilized the results of the *Orf1ab* and *N* gene to reach a conclusion. Successful amplification of both genes and Ct value ≤35 was recognized as positive for SARS-CoV-2 infection; only one site with Ct value ≤35 or both genes between Ct 35 to 38 was taken as inconclusive, and no successful amplification or Ct ≥ 38 was recognized as negative for infection.

### Clinical records of patients

The clinical records of patients were kept in Renmin Hospital of Wuhan University. Clinical, laboratory, and radiological characteristics and treatment and outcome data were obtained using data collection forms from electronic medical records. The data were reviewed by a trained team of physicians. The study and use of all records were approved by the Ethics Committee of Hubei Provincial Renmin Hospital (WDRY2019-K056).

## Data Availability

All data generated or used during the study appear in the submitted article.

## Data availability

All data for support the study result are included in this published article (and its supplementary information files). Other data generated during and/or analyzed during the current study are available from the corresponding author on reasonable request.

